# Diagnostic accuracy of rapid antigen tests in pre-/asymptomatic close contacts of individuals with a confirmed SARS-CoV-2 infection

**DOI:** 10.1101/2021.03.18.21253874

**Authors:** E Schuit, IK Veldhuijzen, RP Venekamp, W van den Bijllaardt, SD Pas, EB Lodder, R Molenkamp, CH GeurtsvanKessel, J. Velzing, RC Huisman, L Brouwer, T Boelsums, GJ Sips, KSM Benschop, L Hooft, JHHM van de Wijgert, S van den Hof, KGM Moons

## Abstract

**Background:** Pre-/asymptomatic close contacts of SARS-CoV-2 infected individuals were tested at day 5 after contact by real-time reverse transcriptase polymerase chain reaction (RT-PCR). Diagnostic accuracy of antigen-detecting rapid diagnostic tests (Ag-RDT) in pre-/asymptomatic close contacts was up till now unknown.

**Methods:** We performed a prospective cross-sectional diagnostic test accuracy study. Close contacts (e.g. selected via the test-and-trace program or contact tracing app) aged ≥16 years and asymptomatic when requesting a test, were included consecutively and tested at day 5 at four Dutch public health service test sites. We evaluated two Ag-RDTs (BD Veritor™ System Ag-RDT (BD), and Roche/SD Biosensor Ag-RDT (SD-B)) with RT-PCR as the reference standard. Virus culture was performed in RT-PCR positive individuals to determine the viral load cut-off above which 95% was culture positive, as a proxy of infectiousness.

**Results:** Of 2,678 BD-tested individuals, 233 (8.7%) were RT-PCR positive and BD detected 149 (sensitivity 63.9%; 95% confidence interval 57.4%-70.1%). Out of 1,596 SD-B-tested individuals, 132 (8.3%) were RT-PCR positive and SD-B detected 83 (sensitivity 62.9%; 54.0%-71.1%). When applying an infectiousness viral load cut-off ≥ 5.2 log10 gene copies/mL, the sensitivity was 90.1% (84.2%-94.4%) for BD, 86.8% (78.1% to 93.0%) for SD-B overall, and 88.1% (80.5%-93.5%) for BD, 85.1% (74.3%-92.6%) for SD-B for those still asymptomatic at the actual time of sampling. Specificity was >99% for both Ag-RDTs in all analyses.

**Conclusions:** The sensitivity for detecting SARS-CoV-2 of both Ag-RDTs in pre-/asymptomatic close contacts is over 60%, increasing to over 85% after applying an infectiousness viral load cut-off.

**Trial registration number:** Not applicable. A study protocol is available upon request.

## Introduction

The cornerstone of COVID-19 epidemic control has been the implementation of generic infection control measures (hand hygiene, physical distancing, and staying at home when symptomatic) combined with test-and-trace programs. Mathematical modelling studies have shown that test-and-trace programs, in combination with generic infection control measures, can successfully control SARS-CoV-2 epidemics, even when assuming that up to 40% of transmissions may occur by pre-/asymptomatic individuals ^1,2^. However, they can only reduce the reproductive number below 1.0 when test-and-trace delays are minimized ^3,4^. In test-and-trace programs, contacts of infected individuals are actively traced and offered testing, initially only when symptomatic, but increasingly also when pre-/asymptomatic ^5^.

In the first phase of the epidemic, testing was performed by reverse transcriptase polymerase chain reaction (RT-PCR) of combined oral-nasal/nasopharyngeal swabs. The sensitivities of these tests increase as the upper respiratory tract viral load increases, and reaches a high plateau on day 5 after infection ^6,7^. While RT-PCR is considered the reference test for SARS-CoV-2, it also has disadvantages. RT-PCR testing platforms are typically only available in centralized laboratories and require sample batching, thereby introducing testing delays. Point-of-care SARS-CoV-2 tests became available during the global pandemic, and of these, lateral flow antigen-detecting rapid diagnostic tests (Ag-RDT) are promising. They require no or minimal equipment, provide a result within minutes, and can be performed in a range of settings with relatively little training.

At the end of 2020, Ag-RDT had been evaluated and considered sufficient to replace RT-PCRs, notably if not only in symptomatic individuals. Diagnostic accuracies may, however, be lower in asymptomatic individuals and in samples containing lower SARS-CoV-2 viral loads ^8^. The latter is not necessarily problematic if lower viral load translates into lower subsequent expected infectiousness ^9^. The very few Ag-RDT evaluations performed in asymptomatic individuals thus far had small sample sizes ^10^, did not take into account whether the tested individual had been exposed to an index case ^11^, nor was virus culture performed ^12^. Therefore, we conducted this first large scale prospective diagnostic test accuracy study in pre-/asymptomatic close contacts of index cases to quantify the accuracy of two Ag-RDTs for detecting SARS-CoV-2 infection with RT-PCR as the reference standard.

## Methods

### Ethical review

The Medical Ethics Review Committee (METC) Utrecht concluded that ethics approval was not required because the study is outside the scope of the Dutch Medical Research Involving Human Subjects Act (protocol number: 20/750). All participants signed an informed consent form prior to any study procedure.

### Study design and population

This prospective cross-sectional diagnostic test accuracy study was embedded within the Dutch routine testing infrastructure. As of 1 December 2020, Dutch policy encourages close contacts who are still pre-/asymptomatic to schedule a test on the fifth day since last exposure to an index case (referred to as a ‘fifth day test’). In the Netherlands, individuals are notified as a close contact by the Dutch public health service test-and-trace program, and/or the Dutch contact tracing mobile phone application (the ‘CoronaMelder’ app) and/or an individual with a confirmed SARS-CoV-2 infection (index case). Participants were recruited consecutively at four Dutch public health service test sites, located in the West-Brabant region (Raamsdonksveer and Roosendaal) and in the city of Rotterdam (Rotterdam Ahoy and Rotterdam The Hague Airport [travellers were not considered]). Close contacts presenting at these test sites were considered eligible if they were aged 16 years or older, scheduled for a fifth day test, asymptomatic at the time of the test request, and willing and able to sign an informed consent in Dutch language.

### Inclusion procedure

Participants arrived at the test sites by car (West-Brabant) or by foot (Rotterdam). Test site personnel approached them and verbally verified study eligibility. Eligible individuals received a study flyer and a participant information letter. After signing the informed consent form, a short questionnaire on presence, type and onset of symptoms (Supplementary Material 1) was completed by participants themselves (West-Brabant) or by test site personnel (Rotterdam), while participants waited for sampling. Questionnaire data were extracted in duplicate by two independent persons.

### Specimen collection, testing procedures and virus culture procedures

A detailed description of collecting, testing, including culturing, of specimens can be found in the Supplementary Material 2. Trained personnel took two combined oropharyngeal-nasal (West-Brabant) or oro-nasopharyngeal (Rotterdam) swabs from each study participant: the first for an RT-PCR test and the second for an Ag-RDT. Swabs were transported to relevant offsite and onsite laboratories, respectively.

During the study period, all study sites were using Roche COBAS6800/8800 platforms for RT-PCR testing (Supplementary Material 2), the sites in West-Brabant were using the BD Veritor™ System for Rapid Detection of SARS-CoV-2 Ag-RDT (‘BD’; Becton, Dickinson and Company, Franklin Lakes, NJ, USA), and the Rotterdam sites the Roche/SD Biosensor SARS-CoV-2 Rapid Antigen Test (‘SD-B’; Roche Diagnostics, Basel, Switzerland). Both Ag-RDT were applied according to manufacturer instructions with one exception: BD results were determined visually instead of using a BD Veritor Plus Analyzer. Interpretation of Ag-RDTs was always done prior to (thus blinded for) RT-PCR. Similarly, Ag-RDT results were not available to those assessing RT-PCR results. Participants received the RT-PCR result, but not the Ag-RDT result, to direct further management (such as quarantaine advice).

At the Erasmus MC Viroscience diagnostic laboratory, samples of participants with a positive RT-PCR test result were cultured for seven days, and, once cytopathic effects (CPE) were visible, the presence of SARS-CoV-2 was confirmed with immunofluorescent detection of SARS-CoV-2 nucleocapsid protein (Rabbit polyclonal antibody, Sino Biological inc.), Eschborn, Germany) (Supplementary Material 2).

### Outcomes and statistical analyses

The primary outcome was the diagnostic accuracy (in terms of sensitivity, specificity, positive and negative predictive values with their 95% confidence intervals) of each Ag-RDT, with RT-PCR as reference standard. Since the number of individuals without RT-PCR or Ag-RDT result was very low (n=21 (0.5%); Figure 1), a complete case analysis was performed. Secondary outcomes included the diagnostic Ag-RDT accuracies stratified for a) the occurrence of COVID-19-like symptoms between test request and time of sampling (yes vs. no), b) the number of days between last contact and date of sampling (<5 vs. 5 vs. >5 days), c) different viral load cut-offs and the viral load cut-off above which 95% of RT-PCR positives had a positive culture as a proxy of infectiousness. To the latter aim, Ct values were first converted into viral loads (virus gene copies/ml) using a standard curve (Supplementary Material 2). The infectiousness cut-off was defined as the viral load above which 95% of RT-PCR positives showed *in vitro* infectivity in cell culture.

**Figure 1.**
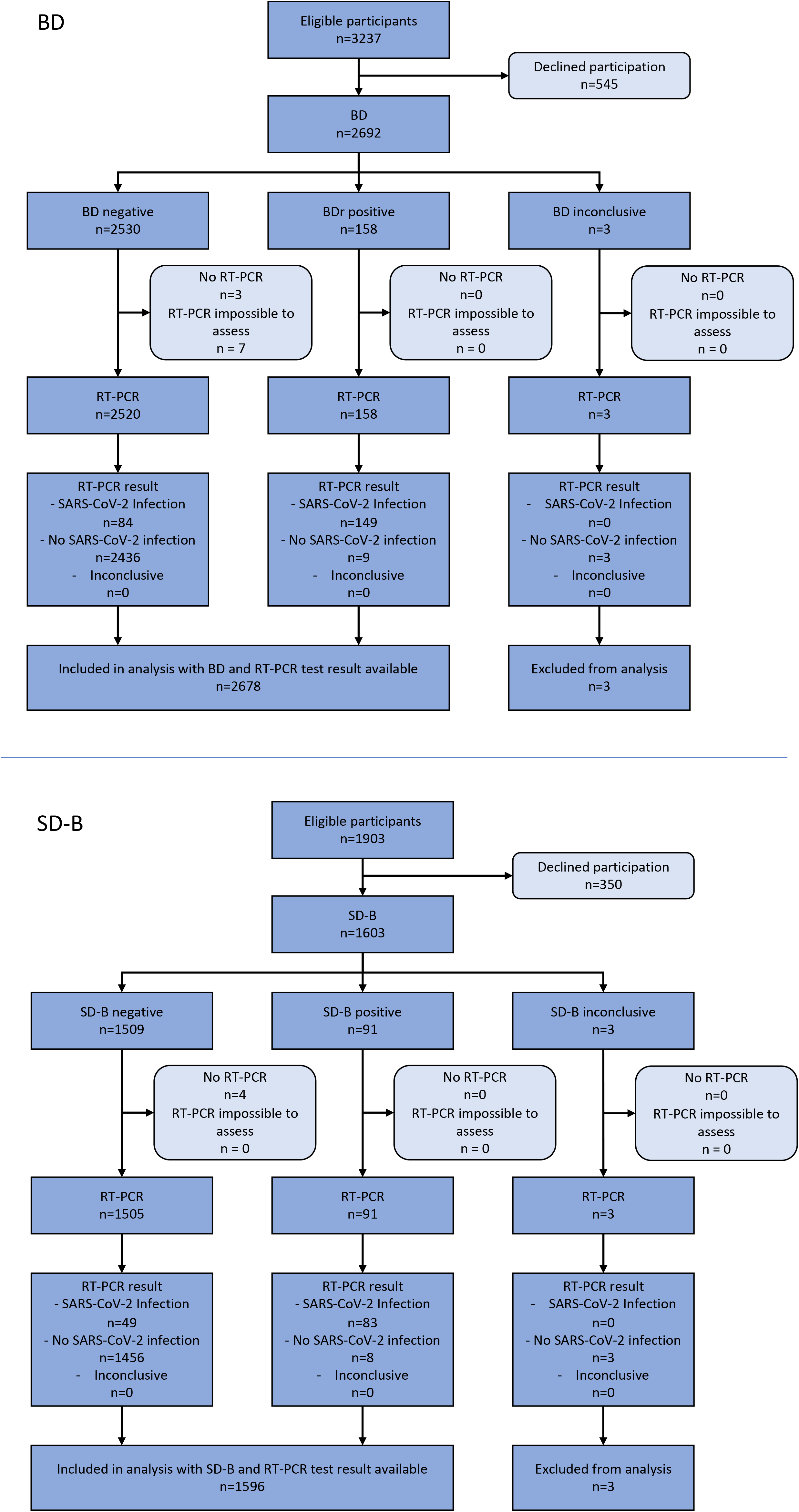
Flow of study participants. BD = BD Veritor™ System for Rapid Detection of SARS-CoV-2 Ag-RDT (‘BD’), SD-B = Roche/SD Biosensor SARS-CoV-2 Rapid Antigen Test.

Finally, we used routine national testing data to determine whether any RT-PCR-negative participants at day 5, afterwards tested positive by RT-PCR or Ag-RDT within 10 days after the initial RT-PCR test, to determine whether testing pre-/asymptomatic individuals at day 5 since last contact with the index case, may be too early.

### Sample size considerations

Previous Ag-RDT performance studies in symptomatic individuals found sensitivities of around 85% ^9,13-15^. We based our sample size calculation on an expected sensitivity of 80%, with a margin of error of 7%, type I error of 5% and power of 90%. Hence, we aimed for 140 positive RT-PCR tests for each Ag-RDT vs. RT-PCR test comparison. We anticipated a SARS-CoV-2 prevalence (based on RT-PCR) in our target population of 10%, and closely monitored RT-PCR test positivity proportion over time to prolong recruitment if needed.

## Results

Between 14 December 2020 and 6 February 2021, 5,191 individuals were considered eligible for participation of whom 4,296 participated (Figure 1). Both RT-PCR and Ag-RDT results were available for 2,678 (99.5%) and 1,596 (99.5%) participants in the BD and SD-B group, respectively. The BD and SD-B groups were similar: respectively the mean ages (standard deviation (SD); in years) were 45.9 (SD 17.6) and 40.7 (SD 16.4), 51.3% and 47.3% were female, and 8.6% and 10.1% had developed symptoms at the time of sampling (Table 1).

**Table 1.** Diagnostic accuracy parameters of both Ag-RDTs

| Analysis |  | # | Prevalence * | Sensitivity [%]<br>(95% CI) | Specificity [%]<br>(95% CI) | Positive Predictive Value [%]<br>(95% CI) | Negative Predictive Value [%]<br>(95% CI) |
| --- | --- | --- | --- | --- | --- | --- | --- |
| ***BD Veritor™ System for Rapid Detection of SARS-CoV-2 Ag-RDT ('BD')*** |  |  |  |  |  |  |  |
| <u>Primary analysis</u> |  | 2678 | 8.7% | 63.9<br>(57.4 to 70.1) | 99.6<br>(99.3 to 99.8) | 94.3<br>(89.5 to 97.4) | 96.7<br>(95.9 to 97.3) |
| <u>Secondary (stratified) analysis</u> |  |  |  |  |  |  |  |
| Infectiousness viral load cut-off<sup>†</sup> |  | 2677<sup>§</sup> | 5.7% | 90.1<br>(84.2 to 94.4) | 99.2<br>(98.8 to 99.5) | 87.3<br>(81.0 to 92.0) | 99.4<br>(99.0 to 99.7) |
| Symptoms at sampling<sup>#</sup> | Yes | 219 | 17.4% | 84.2<br>(68.7 to 94.0) | 99.4<br>(97.0 to 100) | 97.0<br>(84.2 to 99.9) | 96.8<br>(93.1 to 98.8) |
|  | No | 2317 | 7.7% | 58.7<br>(51.1 to 66.0) | 99.6<br>(99.3 to 99.8) | 92.9<br>(86.5 to 96.9) | 96.6<br>(95.8 to 97.4) |
| Interval between sampling and last contact with index case [days]<sup>@</sup> | < 5 | 379 | 14.8% | 69.6<br>(55.9 to 81.2) | 99.7<br>(98.3 to 100) | 97.5<br>(86.8 to 99.9) | 95.0<br>(92.1 to 97.1) |
|  | 5 | 1303 | 6.5% | 62.4<br>(51.2 to 72.6) | 99.9<br>(99.5 to 100) | 98.1<br>(90.1 to 100) | 97.4<br>(96.4 to 98.2) |
|  | > 5 | 511 | 9.0% | 56.5<br>(41.1 to 71.1) | 99.1<br>(97.8 to 99.8) | 86.7<br>(69.3 to 96.2) | 95.8<br>(93.7 to 97.4) |
| ***Roche/SD Biosensor SARS-CoV-2 Rapid Antigen Test ('SD-B')*** |  |  |  |  |  |  |  |
| <u>Primary analysis</u> |  | 1596 | 8.3% | 62.9<br>(54.0 to 71.1) | 99.5<br>(98.9 to 99.8) | 91.2<br>(83.4 to 96.1) | 96.7<br>(95.7 to 97.6) |
| <u>Secondary (stratified) analysis</u> |  |  |  |  |  |  |  |
| Infectiousness viral load cut-off |  | 1596 | 5.7% | 86.8<br>(78.1 to 93.0) | 99.2<br>(98.6 to 99.6) | 86.8<br>(78.1 to 93.0) | 99.2<br>(98.6 to 99.6) |
| Symptoms at sampling<sup>#</sup> | Yes | 158 | 19.0% | 73.3<br>(54.1 to 87.7) | 98.4<br>(94.5 to 99.8) | 91.7<br>(73.0 to 99.0) | 94.0<br>(88.6 to 97.4) |
|  | No | 1414 | 7.1% | 59.4<br>(49.2 to 69.1) | 99.5<br>(99.0 to 99.8) | 90.9<br>(81.3 to 96.6) | 97.0<br>(95.9 to 97.8) |
| Interval between sampling and last contact with index case [days]<br><sup>@</sup> | < 5 | 153 | 13.1% | 75.0<br>(50.9 to 91.3) | 99.2<br>(95.9 to 100) | 93.8<br>(69.8 to 99.8) | 96.4<br>(91.7 to 98.8) |
|  | 5 | 1095 | 7.8% | 61.2<br>(50.0 to 71.6) | 99.5<br>(98.9 to 99.8) | 91.2<br>(80.7 to 97.1) | 96.8<br>(95.6 to 97.8) |
|  | > 5 | 205 | 6.3% | 69.2<br>(38.6 to 90.9) | 99.5<br>(97.1 to 100) | 90.0<br>(55.5 to 99.7) | 97.9<br>(94.8 to 99.4) |

CI = confidence interval; N/A = not applicable.
<sup>\*</sup> SARS-CoV-2 infection based on RT-PCR test result
<sup>#</sup> Symptoms were not available from 142 individuals in the BD group and 24 in the SD-B group
<sup>!</sup> The viral load cut-off for infectiousness, defined as the viral load above which 95% of RT-PCR positives had a positive viral culture, was 5.2 log<sub>10</sub> E gene copies/mL.
<sup>§</sup> Viral load was unavailable for one BD-tested individual with a positive RT-PCR test result
<sup>@</sup> The interval between the moment of sampling and the last contact with an infected individual was not available for 488 individuals in the BD group and 143 individuals in the SD-B group, mainly because this question was added to the questionnaire later in study. Initially, a 3-item questionnaire was used. Questions 1 and 2 in the 5-item questionnaire (see Supplementary Material 1) were added after the first week of the study. In addition, do note that the time interval between the last contact and the time of sampling is not the same as the time between the test request and the time of sampling.

In the BD group, 233 (8.7%) had an RT-PCR-confirmed SARS-CoV-2 infection; 149 were detected by the BD Ag-RDT resulting in an overall sensitivity of 63.9% (95% CI 57.4% to 70.1%) (Table 1). In the SD-B group, 132 (8.3%) had an RT-PCR-confirmed SARS-CoV-2 infection; 83 were detected by the SD-B Ag-RDT resulting in an overall sensitivity of 62.9% (95% CI 54.0% to 71.1%).

In individuals who had developed symptoms between the test request and the time of sampling, the sensitivity was 84.2% (95% CI 68.7% to 94.0%) for BD (N=219) and 73.3% (95% CI 54.1% to 87.7%) for SD-B (N=158). Additional stratified diagnostic accuracy parameters are shown in Table 1. Two by two tables of all primary and secondary analyses are presented in Tables S2 and S3.

Figure 2 shows the distribution of viral loads in individuals with a positive RT-PCR test result, stratified by a combination of the SD-B Ag-RDT result and the ability to culture virus. Above a viral load of 5.2 log10 E-gene copies/mL, 95% of RT-PCR positive individuals showed a positive virus culture. Using that viral load as a cut-off for infectiousness, the sensitivity was 90.1% (95% CI 84.2% to 94.4%) for BD and 86.8% (95% CI 78.1 % to 93.0 %) for SD-B, respectively. Figure 3 shows diagnostic accuracy parameters stratified by different viral load cut-offs. The sensitivity of both Ag-RDTs at the infectious viral load cut-off in persons without symptoms at the time of sampling was 88.1% (95% CI 80.5% to 93.5%) for BD and 85.1% (74.3% to 92.6%). Other diagnostic accuracy parameters for this group are presented in more detail in Table S4, and at varying viral load cut-offs in Supplementary Figure 2.

**Figure 2.**
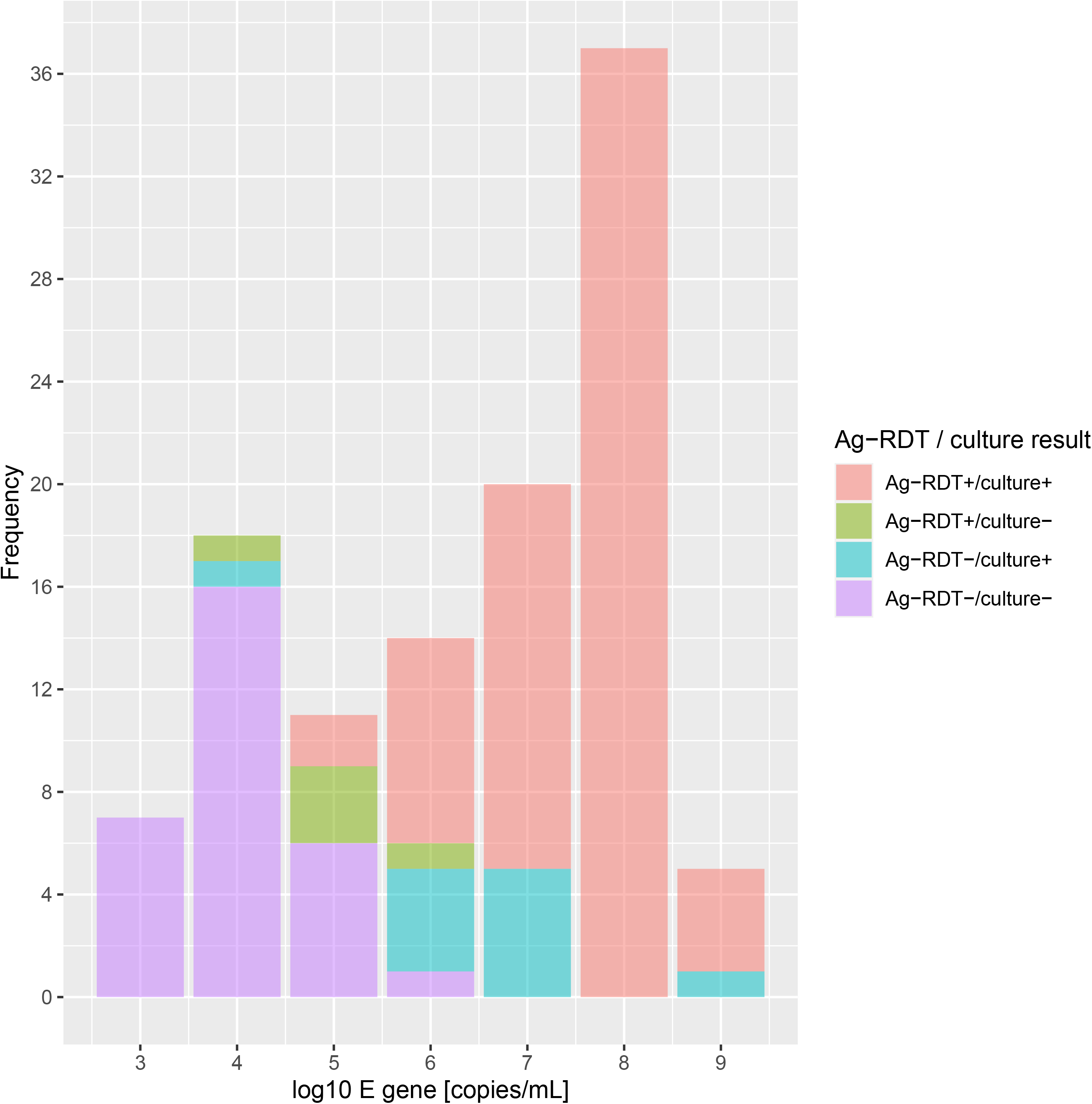
Distribution of viral loads of individuals with a positive RT-PCR test result, stratified by a combination of the Roche/SD Biosensor SARS-CoV-2 Rapid Antigen Test result (Ag-RDT+/-) and the ability to culture (culture +/-), with Ag-RDT+ and Ag-RDT-corresponding to a positive and negative Ag-RDT result, and culture+ and culture-indicating whether it was possible to culture virus or not.

**Figure 3.**
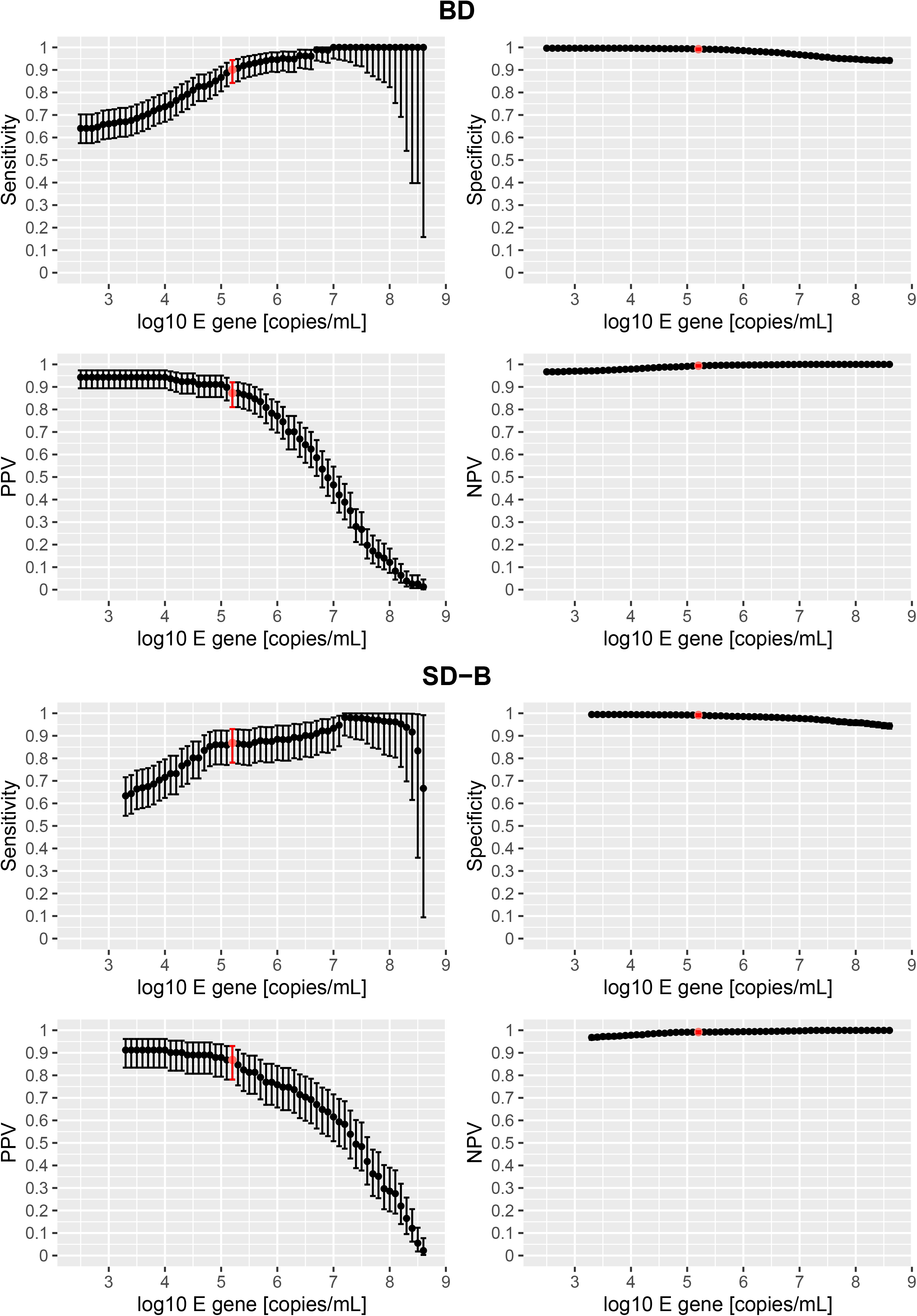
Diagnostic accuracy parameters of both Ag-RDTs for different definitions of RT-PCR test positivity based on viral load cut-offs, where a positive RT-PCR test with a viral load below the viral load cut-off threshold is considered a negative RT-PCR test result. Points highlighted in red indicate a viral load cut-off of 5.2 log10 E gene copies/mL, which was considered the viral load cut-off for infectiousness as determined by viral culture. BD = BD Veritor™ System for Rapid Detection of SARS-CoV-2 Ag-RDT; SD-B = Roche/SD Biosensor SARS-CoV-2 Rapid Antigen Test; PPV = positive predictive value; NPV = negative predictive value.

Finally, routine national testing follow-up information was available for 89% of all included study participants. A total of 31 (1.5%) and 26 (2.0%) individuals had a positive RT-PCR test result within 10 days after an initial negative RT-PCR test result in the BD and SD-B group, respectively.

## Discussion

To our knowledge, this is the largest study thus far to specifically assess the diagnostic test accuracy of Ag-RDTs in pre-/asymptomatic close contacts. Our study shows that two Ag-RDTs, that are routinely used for symptomatic individuals, have around 63% sensitivity for detecting SARS-CoV-2 in pre-/asymptomatic close contacts. However, both Ag-RDTs particularly appeared to show a negative result – while the corresponding RT-PCR test was positive – in those with a lower viral load ranges. At viral loads above 5.2 log10 E gene copies/mL (the assumed viral load cut-off for infectiousness based on viral culture results), both Ag-RDTs showed sensitivities over 85%, i.e. above the WHO sensitivity criteria defined for Ag-RDTs in symptomatic individuals. Specificity was >99% for both Ag-RDTs in all analyses. About 2% of close contacts who initially tested RT-PCR negative at day 5, developed symptoms and subsequently tested positive after day 5. This rather low percentage indicates that testing close contacts that are still asymptomatic at day 5, is not too early.

The prevalence of SARS-CoV-2 infections among all adults, both symptomatic and pre-/asymptomatic, tested as part of contact tracing in all the public health service testing sites in the study period, was 18% ^16^. This percentage is comparable to the prevalence among symptomatic close contacts in our study population. Although the prevalence in pre-/asymptomatic close contacts is expectedly lower, it is considerably higher than the prevalence of 0.8%-1% (at the time of the study period) as estimated in the general population of the Netherlands ^16^, and in other countries ^17,18^.

Only about 2% of close contacts who initially tested RT-PCR negative at day 5 after the close contact developed symptoms and subsequently tested positive, i.e. infections missed by RT-PCR on day 5 after exposure to an index case. These proportions of ‘missed’ infections are expected to increase when using an Ag-RDT instead of RT-PCR. This underlines the importance of immediate self-isolation and repeat testing when symptoms develop after a negative day 5 Ag-RDT.

The extent to which the lower sensitivity of Ag-RDTs compared to RT-PCR outweighs the positive effects of simplified logistics, reduced delays, and the potential for self-testing, is currently unknown. We will monitor this utiziling national test-and-trace information. Modeling studies will also help to further address this question in the future.

We were also able to minimize missing values and measurement bias by taking samples for the Ag-RDT and RT-PCR from each participant at the same time, using the same Ag-RDT and RT-PCR reference test for all participants, and having all (index and reference) tests performed by trained personnel who were blinded to the result of the other test.

Our study also has some potential limitations. First, although we aimed to sample pre-/asymptomatic individuals at five days after exposure to an index case, 12% was sampled before the fifth day since last contact. It is known that the accuracy of the RT-PCR reference test is not yet optimal before the fifth day after contact with an index case ^6^. Interestingly though, our stratified analysis indicated that the RT-PCR positivity fraction was actually higher in close contacts who were sampled before the fifth day since last contact. We hypothesize that some of these individuals have had prolonged contact with the index case, for example because they live in the same household. Close contacts living in the same household on average test more often positive compared to non-household close contacts (20% vs. 10%) ^16^. This would also explain why these close contacts reported to have contact with the index case less than 5 days ago, as the last contact with an infected household member could be the same day of testing. A second limitation is that virus culture was only available in one of the two central laboratories. Hence, the assumed infectiousness viral load cut-off was extrapolated to the second laboratory. Reassuringly, the RT-PCR calibration curves of both laboratories indicated that Ct values corresponded to similar viral loads in both laboratories. A correlation between infectivity in culture and viral load of the specimen as well as negative associations between lower viral loads and secondary attack rates have been established ^19-22^. Some uncertainty remains as the exact upper respiratory tract viral load cut-off below which transmissions no longer take place is yet unknown, and due to high variability in methodologies and results between laboratories and studies ^21,22^.

The Dutch Outbreak Management Team (OMT) that provides guidance to the Ministry of Health, Welfare and Sport on policy regarding COVID-19, advised, based on the results of this study, that pre-/asymptomatic close contacts of individuals with a confirmed SARS-CoV-2 infection can be tested for SARS-CoV-2 using an Ag-RDT. As a result the Dutch policy now allows testing of close contacts using Ag-RDTs from day 5 onwards, even when they have not (yet) developed symptoms. Accordingly, positive test results are known and communicated earlier such that the use of Ag-RDTs in pre-/asymptomatic close contacts has the potential to help prevent onward SARS-CoV-2 transmission.

## Supporting information

STARD reporting guideline

## Data Availability

Data will not available for sharing.

## Acknowledgements

We would like to acknowledge all participants, study personnel at the local public health service test sites, personnel at the participating labs, and study personnel at the RIVM that helped process the questionnaires, design and distribute the study forms. A special thanks goes to Esther Stiefelhagen, Roel Ensing, and Wendy Mouthaan for their efforts in the logistics towards and at the local test sites. Written permission was obtained from all three to include their names in this Acknowledgment section. ES, RE, and WH did not receive any compensation for their contributions.

## Author contributions

KGMM and JHHMvdW initiated the study. ES, IKV, RPV, WvdB, EL, RM, GJS, KB, LH, JHHMvdW, SvdH, and KGMM designed the study. IKV coordinated the study. WvdB, SDP, RM, JV, and RCH were responsible for lab analyses and data processing. CHGvK performed virus culture. ES performed the statistical analysis in close collaboration with IKV and KGMM. ES, IKV, RPV, SvdH, and KGMM drafted the first version of the manuscript. All authors critically read the manuscript and provided feedback. All authors approved the submission of the current version of the manuscript.

## Additional information

(containing supplementary information line (if any) and corresponding author line).

The study protocol is available upon request by contacting Karel Moons at.

