## supplement for "Diagnostic accuracy of rapid antigen tests in pre-/asymptomatic close contacts of individuals with a confirmed SARS-CoV-2 infection"

#### Table of contents

**Table S1** Baseline characteristics of pre-/asymptomatic close contacts of individuals with a confirmed SARS-CoV-2 infection. 2

**Table S2** Two-by-two tables used in primary and secondary analysis to determine diagnostic accuracy parameters of the BD Veritor$^{TM}$ System for Rapid Detection of SARS-CoV-2 Ag-RDT ('BD') 3

**Table S3** Two-by-two tables used in primary and secondary analysis to determine diagnostic accuracy parameters of the Roche/SD Biosensor SARS-CoV-2 Rapid Antigen Test ('SD-B') 4

**Table S4** Diagnostic accuracy parameters of both Ag-RDTs in asymptomatic close contacts, i.e., without symptoms, at sampling at the viral load cut-off for infectiousness at 5·2 log10 E gene copies/mL, the viral load above which 95% of RT-PCR positives had a positive culture. 5

**Supplementary Figure 1** Diagnostic accuracy parameters of Ag-RDTs in asymptomatic close contacts, i.e., without symptoms at sampling for different viral load cut-offs. 6

**Supplementary Material 1:** short questionnaire (translated from Dutch) 7
3-item questionnaire 7
5-item questionnaire 8

**Supplementary Material 2:** Specimen collection, SARS-CoV-2 diagnostic testing, and SARS-CoV-2 virus culture procedures 9

**References** 10

**Table S1 Baseline characteristics of pre-/asymptomatic close contacts of individuals with a confirmed SARS-CoV-2 infection.**

| | Stratified by Ag-RDT | |
|---|---|---|
| | BD | SD-B |
| | N = 2678 | N = 1596 |
| Age [years], mean (SD) | 45·9 (17·6) | 40·7 (16·4) |
| Gender, female n (%) | 1370 (51·3) | 751 (47·3) |
| Time interval between last contact and sampling [days], median (IQR), range (min-max) | 5 (5 to 5), (0 to 13) | 5 (5 to 5), (0 to 11) |
| Symptoms at time of sampling, n (%) | 219 (8·6) | 158 (10·1) |
| Symptom onset, n (%)* | *N = 219* | *N = 158* |
| At day of sampling | 17 (7·8) | 14 (8·9) |
| A day before sampling | 64 (29·2) | 37 (23·4) |
| Two days before sampling | 51 (23·3) | 39 (24·7) |
| Three or more days before sampling | 83 (37·9) | 45 (28·5) |
| Unknown | 4 (1·8) | 23 (14·6) |
| Type of symptoms (self-reported), n (%)*# | *N = 219* | *N = 158* |
| Common cold | 167 (76·3) | 123 (77·8) |
| Shortness of breath | 25 (11·4) | 12 (7·6) |
| Fever | 13 (5·9) | 9 (5·7) |
| Coughing | 60 (27·4) | 24 (15·2) |
| Loss of taste or smell | 6 (2·7) | 5 (3·2) |
| Muscle ache | 18 (8·2) | 5 (3·2) |
| Other symptoms | 16 (7·3) | 15 (9·5) |

BD = BD Veritor™ System for Rapid Detection of SARS-CoV-2 Ag-RDT; IQR = inter quartile range; min=minimum; max=maximum; SD=standard deviation; SD-B = Roche/SD Biosensor SARS-CoV-2 Rapid Antigen Test.

In the Netherlands, individuals are notified of a close contact by the Dutch public health service test-and-trace program, and/or the Dutch contact tracing mobile phone application (the CoronaMelder app) and/or an individual with a confirmed SARS-CoV-2 infection (index case).

* percentage calculated as proportion of those with symptoms at time of sampling

### totals add up to a number higher than the number of individuals with symptoms at the time of sampling because individuals could report more than one symptom.

**Table S2 Two-by-two tables used in primary and secondary analysis to determine diagnostic accuracy parameters of the BD Veritor$^{TM}$ System for Rapid Detection of SARS-CoV-2 Ag-RDT ('BD')**

| | | | | | |
|---|---|---|---|---|---|
| Primary analysis | | | RT-PCR + | RT-PCR - | Total |
| | | BD test + | 149 | 9 | 158 |
| | | BD test - | 84 | 2436 | 2520 |
| | | Total | 233 | 2445 | 2678 |
| Secondary (stratified) analysis | | | | | |
| Infectiousness viral load cut-off$ | | | RT-PCR + | RT-PCR - | Total |
| | | BD test + | 137 | 20 | 157 |
| | | BD test - | 15 | 2505 | 2520 |
| | | Total | 152 | 2525 | 2677 |
| Symptoms at sampling# | Yes | | RT-PCR + | RT-PCR - | Total |
| | | BD test + | 32 | 1 | 33 |
| | | BD test - | 6 | 180 | 186 |
| | | Total | 38 | 181 | 219 |
| | No | | RT-PCR + | RT-PCR - | Total |
| | | BD test + | 105 | 8 | 113 |
| | | BD test - | 74 | 2130 | 2204 |
| | | Total | 179 | 2138 | 2317 |
| Interval between sampling and last contact with index case [days]@ | < 5 | | RT-PCR + | RT-PCR - | Total |
| | | BD test + | 39 | 1 | 40 |
| | | BD test - | 17 | 322 | 339 |
| | | Total | 56 | 323 | 379 |
| | 5 | | RT-PCR + | RT-PCR - | Total |
| | | BD test + | 53 | 1 | 54 |
| | | BD test - | 32 | 1217 | 1249 |
| | | Total | 85 | 1218 | 1303 |
| | > 5 | | RT-PCR + | RT-PCR - | Total |
| | | BD test + | 26 | 4 | 30 |
| | | BD test - | 20 | 461 | 481 |
| | | Total | 46 | 465 | 511 |

### Symptoms were not available from 142 individuals

$ The viral load cut-off for infectiousness, defined as the viral load above which 95% of RT-PCR positives had a positive culture, was 5·2 log10 E gene copies/mL.

@ The interval between the moment of sampling and the last contact with an index case was not available for 488 individuals, mainly because this question was added to the questionnaire later in study.

**Table S3 Two-by-two tables used in primary and secondary analysis to determine diagnostic accuracy parameters of the Roche/SD Biosensor SARS-CoV-2 Rapid Antigen Test ('SD-B')**

| | | | | | |
|---|---|---|---|---|---|
| Primary analysis | | | RT-PCR + | RT-PCR - | Total |
| | | SD-B test + | 83 | 8 | 91 |
| | | SD-B test - | 49 | 1456 | 1505 |
| | | Total | 132 | 1464 | 1596 |
| Secondary (stratified) analysis | | | | | |
| Infectiousness viral load cut-off[$] | | | RT-PCR + | RT-PCR - | Total |
| | | SD-B test + | 79 | 12 | 91 |
| | | SD-B test - | 12 | 1493 | 1505 |
| | | Total | 91 | 1505 | 1596 |
| Symptoms at sampling[#] | Yes | | RT-PCR + | RT-PCR - | Total |
| | | SD-B test + | 22 | 2 | 24 |
| | | SD-B test - | 8 | 126 | 134 |
| | | Total | 30 | 128 | 159 |
| | No | | RT-PCR + | RT-PCR - | Total |
| | | SD-B test + | 60 | 6 | 66 |
| | | SD-B test - | 41 | 1307 | 1348 |
| | | Total | 101 | 1313 | 1414 |
| Interval between sampling and last contact with index case [days][@] | < 5 | | RT-PCR + | RT-PCR - | Total |
| | | SD-B test + | 15 | 1 | 16 |
| | | SD-B test - | 5 | 132 | 137 |
| | | Total | 20 | 133 | 153 |
| | 5 | | RT-PCR + | RT-PCR - | Total |
| | | SD-B test + | 52 | 5 | 57 |
| | | SD-B test - | 33 | 1005 | 1038 |
| | | Total | 85 | 1010 | 1095 |
| | > 5 | | RT-PCR + | RT-PCR - | Total |
| | | SD-B test + | 9 | 1 | 10 |
| | | SD-B test - | 4 | 191 | 195 |
| | | Total | 13 | 192 | 205 |

[#] Symptoms were not available from 24 individuals
[$] The infectiousness viral load cut-off, defined as the viral load above which 95% of RT-PCR positives had a positive culture, was 5·2 log10 E gene copies/mL.
[@] The interval between the moment of sampling and the last contact with an index case was not available for 143 individuals, mainly because this question was added to the questionnaire later in study.

**Table S4 Diagnostic accuracy parameters of both Ag-RDTs in asymptomatic close contacts, i.e., without symptoms, at sampling at the viral load cut-off for infectiousness at 5·2 log10 E gene copies/mL, the viral load above which 95% of RT-PCR positives had a positive culture**

| Number of asymptomatic close contacts at time of sampling | Prevalence* | Sensitivity [%] (95% CI) | Specificity [%] (95% CI) | Positive Predictive Value [%] (95% CI) | Negative Predictive Value [%] (95% CI) |
|---|---|---|---|---|---|
| ***BD Veritor™ System for Rapid Detection of SARS-CoV-2 Ag-RDT*** | | | | | |
| 2317 | 4·7% | 88·1<br>(80·5 to 93·5) | 99·2<br>(98·8 to 99·6) | 85·0<br>(77·0 to 91·0) | 99·4<br>(99·0 to 99·7) |
| ***Roche/SD Biosensor SARS-CoV-2 Rapid Antigen Test ('SD-B')*** | | | | | |
| 1414 | 4·7% | 85·1<br>(74·3 to 92·6) | 99·3<br>(98·7 to 99·7) | 86·4<br>(75·7 to 93·6) | 99·3<br>(98·6 to 99·6) |

CI = confidence interval; N/A = not applicable

* SARS-CoV-2 infection based on RT-PCR test result

**Supplementary Figure 1 Diagnostic accuracy parameters of Ag-RDTs in asymptomatic close contacts, i.e., without symptoms at sampling for different viral load cut-offs. Points highlighted in red indicate a viral load cut-off of 5.2 log10 E gene copies/mL, which was considered the infectiousness viral load cut-off determined by viral culture. BD = BD Veritor™ System for Rapid Detection of SARS-CoV-2 Ag-RDT; SD-B = Roche/SD Biosensor SARS-CoV-2 Rapid Antigen Test; PPV = positive predictive value; NPV = negative predictive value.**

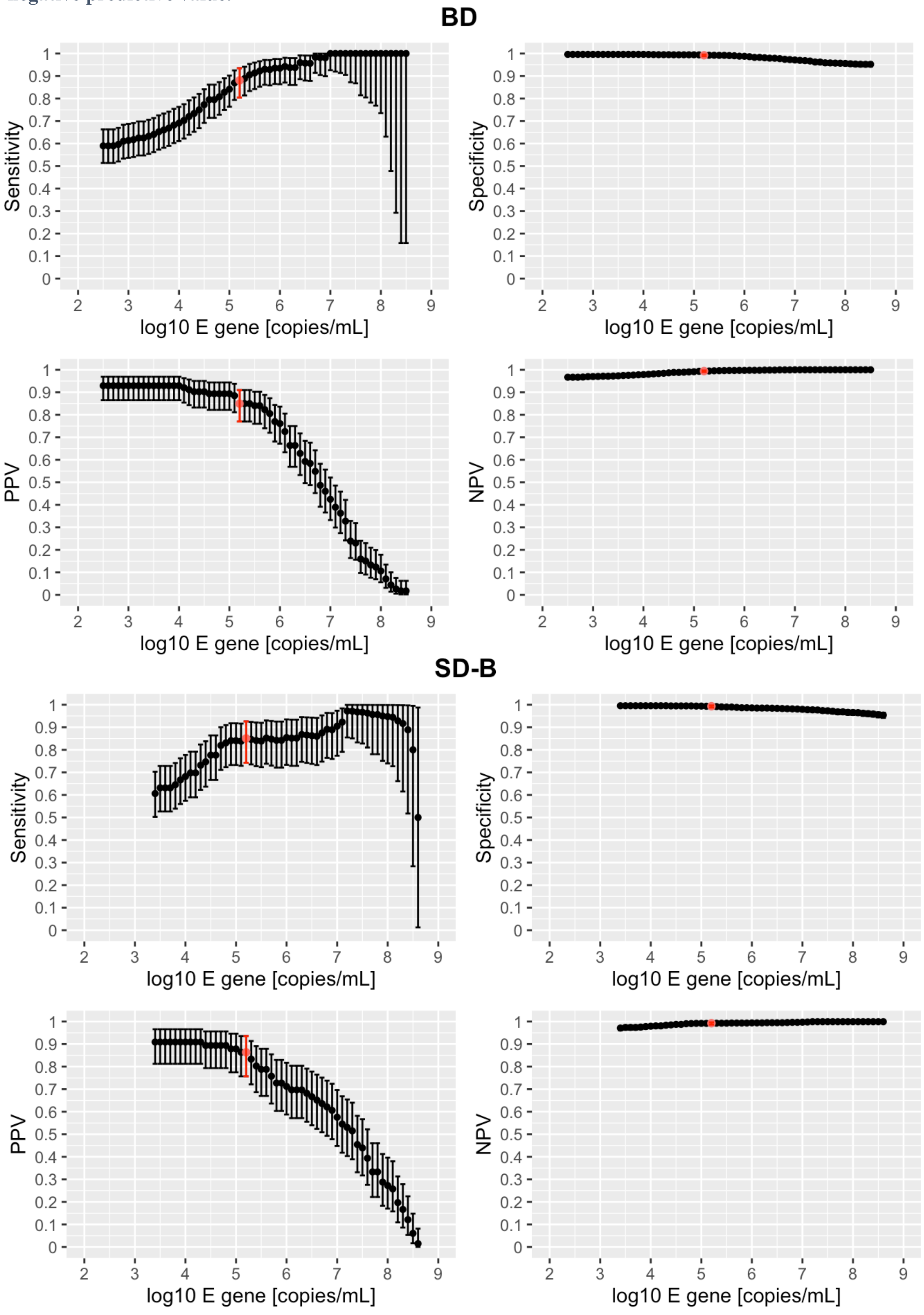

#### Supplementary Material 1: short questionnaire (translated from Dutch)

**3-item questionnaire used between 14 December and 19 December 2020 (West-Brabant region) and 15 December and 18 December 2020 (city of Rotterdam).**

**Short questionnaire on COVID-19 like symptoms and reason for testing**

1. At this moment, do you have any COVID-19 like symptoms?
   - ☐ No *END OF QUESTIONNAIRE*
   - ☐ Yes

2. What COVID-19 like symptoms do you currently have?
   *Multiple answers possible*
   - ☐ Common cold
   - ☐ Shortness of breath
   - ☐ Fever
   - ☐ Coughing
   - ☐ Loss of taste or smell
   - ☐ Muscle ache
   - ☐ I have other symptoms

3. What was the moment you first experienced these symptoms?
   - ☐ Today
   - ☐ Yesterday
   - ☐ Two days ago
   - ☐ Three or more days ago

**5-item questionnaire used from December 19 (city of Rotterdam) and 20 (West-Brabant region) onwards**

**Short questionnaire on COVID-19 like symptoms and reason for testing**

1. What is the reason for testing?
   *Multiple answers possible*
   ☐ Received notification by public health service (by phone or letter)
   ☐ Received notification by CoronaMelder app (English: Corona notification app)
   ☐ Received notification by SARS-CoV-2 infected person
   ☐ None of the above, requested test because of a SARS-CoV-2 infected person in my immediate surroundings

2. When was your last contact with the infected person?
   Date: ___ - ___ - 20___ *(day – month – year)*

3. At this moment, do you have any COVID-19 like symptoms?
   ☐ No *END OF QUESTIONNAIRE*
   ☐ Yes

4. What COVID-19 like symptoms do you currently have?
   *Multiple answers possible*
   ☐ Common cold
   ☐ Shortness of breath
   ☐ Fever
   ☐ Coughing
   ☐ Loss of taste or smell
   ☐ Muscle ache
   ☐ I have other symptoms

5. What was the moment you first experienced these symptoms?
   ☐ Today
   ☐ Yesterday
   ☐ Two days ago
   ☐ Three or more days ago

#### Supplementary Material 2: Specimen collection, SARS-CoV-2 diagnostic testing, and SARS-CoV-2 virus culture procedures

##### Specimen collection and SARS-CoV-2 diagnostic testing procedures

*West-Brabant: RT-PCR and BD Ag-RDT*

Two swabs were taken per participant. The first swab was a combined oropharyngeal- and nasal swab (2·5 cm deep from the edge of the nostril) that was placed in universal transport medium (HiViralTM) with MagnaPure LC lysis- and binding buffer (LBB) (Roche Diagnostics Netherlands, Almere, The Netherlands) and transported to Microvida location Roosendaal laboratory for RT-PCR. RT-PCR was performed using the cobas® SARS-CoV-2 test on the cobas® 8800 platform (Roche Diagnostics International, Rotkreuz, Switzerland). This RT-PCR test has two targets: The E-gene and RdRp-gene. The viral load in genome copies/ml was calculated based on an in-house established standard curve. The second swab was a combined oropharyngeal- and nasal swab (2·5 cm deep) and was placed in a sterile dry tube and frozen at -20°C within 30 minutes after collection before transportation to the Microvida location Amphia laboratory. There, after allowing the specimen to thaw, a trained laboratory technician performed the BD Ag-RDT (BD Veritor System for Rapid Detection of SARS-CoV-2, Becton, Dickinson and Company, Franklin Lakes, NJ, USA) in accordance with the manufacturer's operating procedure within 6 hours after the specimen was obtained. The BD Ag-RDT is a chromatographic immunoassay intended for the direct and qualitative detection of SARS-CoV-2 nucleocapsid antigens in nasal swabs from individuals who are suspected of COVID-19 within the first 5 days of symptom onset. The system is intended to be used with a digital reader although validated for visual reading [1]; we used visual reading, all results were confirmed by a second person.

*Rotterdam: RT-PCR and SD-B Ag-RDT*

Two swabs were taken per participant. First, one combined oro- and nasopharyngeal swab (>5 cm deep from the edge of the nostril) was taken for RT-PCR, placed directly in universal transport media (HiViralTM) and shipped to the Erasmus MC Viroscience diagnostic laboratory. Routine RT-PCR testing was performed on the combined oro- and nasopharyngeal swab in virus transport medium using the cobas ® SARS-CoV-2 test on the cobas 6800® platform (Roche Diagnostics International, Rotkreuz, Switzerland). Genome copies/ml was calculated based on an in house established standard curve. The virus transport medium from the same oro- and nasopharyngeal swab was also directly inoculated onto Vero cells clone 118, without prior freezing [2].

A second nasopharyngeal swab (>5 cm deep from the edge of the nostril) was taken subsequently from the same nostril using the swab included in the kits for the SD-B Ag-RDT (Roche/SD Biosensor SARS-CoV-2 Rapid Antigen Test, Basel, Switzerland). This test was carried out immediately on-site following manufacturer's instructions. Interpretation and recording of test results was performed independently by two persons according to the manufacturer's instructions.

##### SARS-CoV-2 virus culture

At the Erasmus MC Viroscience diagnostic laboratory, samples of participants with a positive RT-PCR test result were inoculated onto Vero cells, and incubated for seven days. Once cytopathic effects were visible, the presence of SARS-CoV-2 virus was confirmed with immunofluorescent detection of its nucleocapsid protein (Rabbit polyclonal antibody Sino Biological inc., Eschborn, Germany).

##### Viral load calculation

All Ct values were converted into viral loads (virus gene copies/ml). The two RT-PCR labs tested the same RIVM SARS-CoV-2 viral load panel, which showed that the Ct values generated in the two laboratories corresponded well (p-value for difference 0·29). Because SARS-CoV-2 virus culture was available in the Erasmus MC only, the Erasmus MC in house RT-PCR viral load standard curve was used for standardization of Ct-viral load conversions in the two laboratories. To account for the lower volume of medium used per swab by Microvida (0·9 mL) than in Erasmus MC (3 mL), the viral loads inferred from Ct values in Microvida were divided by a factor 3·3. The infectiousness viral load cut-off was defined as the viral load above which 95% of RT-PCR positives showed *in vitro* infectivity in cell culture.
